# A practical workflow for organizing clinical intraoperative and long-term iEEG data in BIDS

**DOI:** 10.1101/2020.12.07.20245290

**Authors:** Matteo Demuru, Dorien van Blooijs, Willemiek Zweiphenning, Dora Hermes, Frans Leijten, Maeike Zijlmans, on behalf of the RESPect group

## Abstract

The neuroscience community increasingly uses the Brain Imaging Data Structure (BIDS) to organize data, extending from MRI to electrophysiology data. While automated tools and workflows are developed that help organize MRI data from the scanner to BIDS, these workflows are lacking for clinical intracranial EEG (iEEG data). We present a practical guideline on how to organize full clinical iEEG epilepsy data into BIDS. We present electrophysiological datasets recorded from twelve subjects who underwent intracranial monitoring followed by resective epilepsy surgery at the University Medical Center Utrecht, the Netherlands, and became seizure-free after surgery. These data include intraoperative electrocorticography recordings from six patients, long-term electrocorticography recordings from three patients and stereo-encephalography recordings from three patients. We describe the 6 steps in the pipeline that are essential to structure the data from these clinical iEEG recordings into BIDS and the challenges during this process. These guidelines enable centers performing clinical iEEG recordings to structure their data to improve accessibility, reusability and interoperability of clinical data.

**Background & Summary:** Today’s era of big data and open science has highlighted the importance of organizing and storing data in keeping with the FAIR Data Principles of Findable, Accessible, Interoperable and Reusable Data to the neuroscientific community^1,2^. Over the past five years, a community-driven effort to develop a simple standardized method of organizing, annotating and describing neuroimaging data has resulted in the Brain Imaging Data Structure (BIDS). BIDS was originally developed for magnetic resonance imaging data (MRI^3^), but now also has extensions for magnetoencephalography (MEG^4^), electroencephalography (EEG^5^), and intracranial encephalography (iEEG^6^). BIDS prescribes rules about the organization of the data itself, with a formalized file/folder structure and naming conventions, and provides standardized templates to store associated metadata in human and machine readable, text-based, JSON and TSV file formats. Software packages analyzing neuroimaging data increasingly support data organized using the BIDS format (https://bids-apps.neuroimaging.io/apps/). However, a major challenge in the use of BIDS is to curate the data from their source format into a BIDS validated set. Several tools exist to convert MRI source data into BIDS datasets^7–11^, but to our knowledge, there is currently no tool or protocol for iEEG.

The University Medical Center in Utrecht, the Netherlands, is a tertiary referral center performing around 150 epilepsy surgeries per year. The success of surgery for treating focal epilepsy depends on accurate prediction of brain tissue that needs to be removed or disconnected to yield full seizure control. People referred for epilepsy surgery undergo an extensive presurgical work-up, starting with MRI and video-EEG and, if needed, PET or ictal SPECT. This noninvasive phase is followed directly by a resection, possibly guided by intraoperative ECoG, or by long-term electrocorticography (ECoG) or stereo-encephalography (SEEG) with electrodes placed on or implanted in the brain^12^. From January 2008 until December 2019, 560 of the epilepsy surgeries in our center were guided by intraoperative ECoG; 163 surgeries followed after long-term ECoG or SEEG investigation. These iEEG data offer a unique combination of high spatial and temporal resolution measurements of the living human brain and it is important to curate these data in a way such that they can be used by many people in the future to study epilepsy and typical brain dynamics.

As part of RESPect (Registry for Epilepsy Surgery Patients, ethical committee approval (18-109)), we started to retrospectively convert raw, unprocessed, clinical iEEG data of patients that underwent epilepsy surgery from January 2008 onwards, to the iEEG-BIDS format and identified 6 critical steps in this process. With this paper, we give a practical workflow of how we collected iEEG data in the UMC Utrecht and converted these data to BIDS. We share our entire pipeline and provide practical examples of six patients with intraoperative ECoG, three patients with long-term ECoG and three patients with SEEG data, demonstrating how BIDS can be used for intraoperative as well as long-term recordings.

## Methods

### Patients

Patients who underwent epilepsy surgery in the UMC Utrecht from 2008 onwards are included in RESPect, the Registry for Epilepsy Surgery Patients. For patients operated between January 2008 and December 2017, the medical research ethical committee waived the need to ask for informed consent, so those patients were directly included. Since January 2018, we explicitly ask patients informed consent to collect their data for research purposes.

### iEEG data

Organizing data in BIDS requires a logical grouping of study data into sessions, runs and tasks. We describe the workflow for three different types of iEEG data collected: intraoperative ECoG data collected during surgery, long-term ECoG data and long-term SEEG data collected during several days of epilepsy monitoring.

#### Intraoperative ECoG

Intraoperative ECoG can be performed during epilepsy and tumor surgeries to map brain function or interictal epileptiform activity. In the UMC Utrecht, intraoperative ECoG is performed in lesional epilepsy cases with concordant results of non-invasive examinations, to determine the extent of the neocortical resection, and/or the involvement of mesiotemporal structures and necessity of a hippocampectomy. It usually involves a lesionectomy and possibly a corticectomy of the surrounding tissue based on ECoG findings. It requires careful analysis of pattern, morphology, frequency and localization of interictal activity recorded directly from the exposed cortical surface, in the operating room. Over time, the clinical neurophysiologists in our center developed a standardized procedure of how to perform intraoperative ECoG recordings to tailor epilepsy surgery. Surgery with intraoperative ECoG is composed of three main situations that can be logically grouped into BIDS sessions:

- Pre-resection sessions, consisting of all recordings (with different configurations of the grid and strips/depth) carried out before the surgeon has started the planned resection (see Figure 1A; situation 1A to 1D).
- Intermediate sessions, consisting of all subsequent recordings performed before any iterative extension of the resection area (see Figure 1A; situation 2A to 2D).
- Post-resection sessions, consisting of all the recordings performed after the last resection (see Figure 1A; situation 3A).

**Figure 1.**
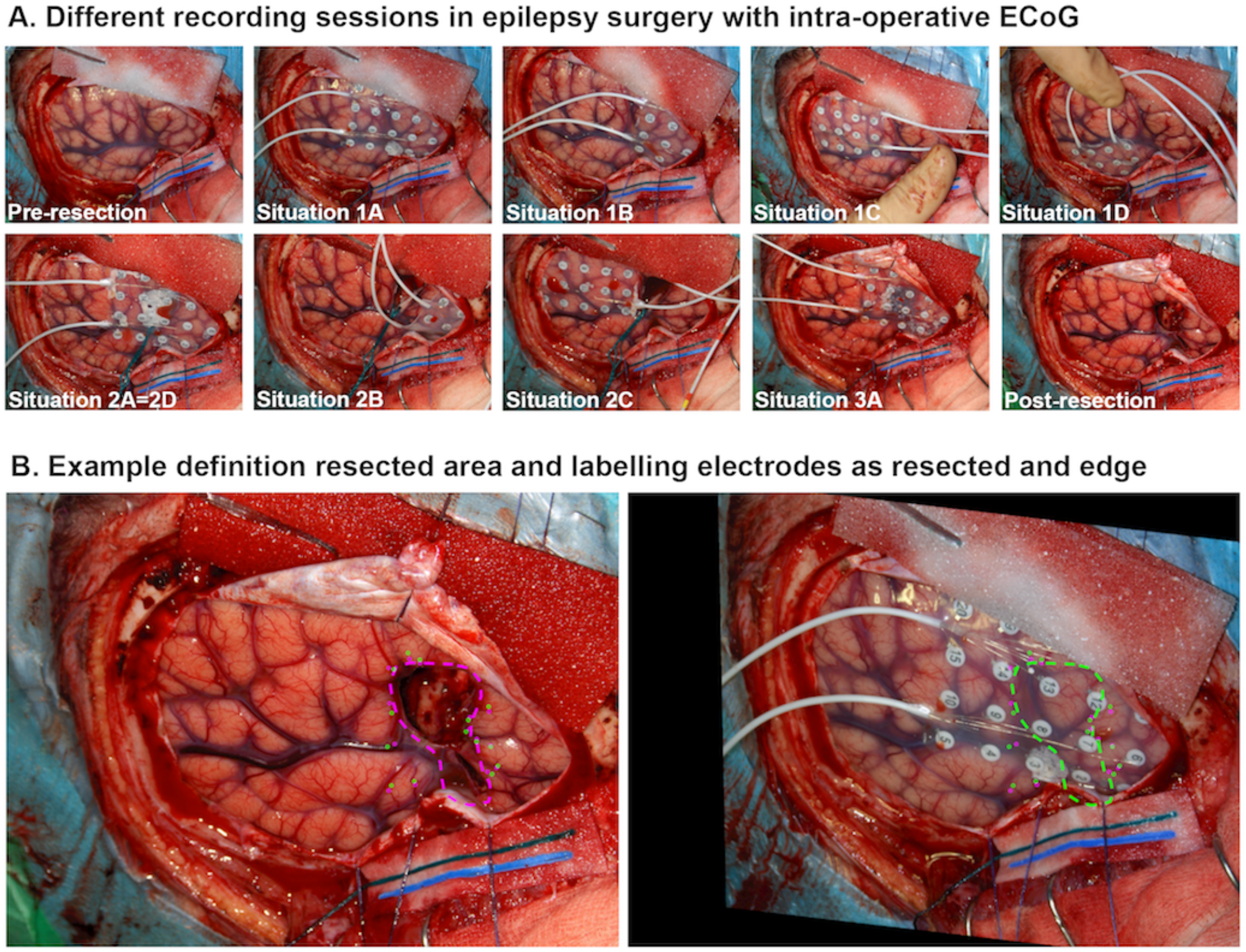
Patient example of the different situations composing a surgery with intraoperative ECoG (A) and how the resected and edge electrodes are defined (B). A) Patient RESP0384 had nine situations recorded. Four situations consist of the pre-resection recordings, and are grouped under BIDS session 1A-D; four situations are recorded during intermediate periods, and are grouped under session 2A-D; there is one post-resection situation, session 3A. B) We used a custom made-software^14^ to align the pre-resection and intermediate session pictures with the post-resection picture. Then we drew the resection area on the post-resection picture and this was automatically projected on the pre-resection and intermediate session pictures (green dashed line). Electrodes that were completely or partly (so exactly on the edge) on top of the resected area were defined as resected. Electrodes that were partly on top of the resected area (so exactly on the edge) or within 0.5 cm of the edge of the resection were defined as edge.

Before each situation a photo is taken to keep track of the grid and strip/depth electrode positions (see Figure 1A). Each situation is labelled with an increasing number starting from 1 (indicative of the period in time respective to the surgical resection) and a consecutive letter starting from A (indicative of the position of the grid and strip/depth for a given session), see example in Figure 1A. Please note that there can be different rounds of intermediate recordings followed by resections if there is still epileptic activity present in the intermediate recording. The recording after the last resection is the final post-resection session and has the highest number. This logical grouping allowed us to store the data in BIDS across sessions.

#### Long-term iEEG

Long-term iEEG recording is performed if results of non-invasive examinations are discordant, but one or more focal hypotheses can be formulated to explain the patient’s seizure manifestations, or if the presumed epileptogenic zone is in or close to the eloquent cortex. Patients are implanted with ECoG or SEEG electrodes placed in locations that will help confirm or rule out the pre-surgical hypothesis based on the results of non-invasive examinations. After implantation of the electrodes, the patient is taken from the operating room to the invasive epilepsy monitoring unit where simultaneous video and intracranial brain signals are recorded for 5-14 days, depending on seizure frequency, type of implantation and clinical performance. During this period, seizures are recorded and functional testing and cortical mapping is performed. The goal of long-term iEEG is to define the volume of cortical tissue generating interictal epileptiform discharges, pinpoint exactly where the seizures start, and ‘map’ the brain tissue surrounding the presumed epileptogenic focus to identify functional tissue that may be impacted by a possible resection. If the epileptogenic focus can be localized, and a surgical strategy can be proposed, the removal of electrodes is followed by a resection. In patients implanted with ECoG electrodes, this resection often takes place in the same surgery as the electrode explantation. Patients implanted with SEEG electrodes do not need a surgery to remove the electrodes, so in these patients the resection is planned in a separate surgery.

In long-term recordings, data recorded within one monitoring period, are logically grouped in the same BIDS session and stored across runs indicating the day and time point of recording in the monitoring period.

### Recording devices for iEEG

IEEG data were recorded using different Micromed headboxes (MicroMed, Mogliano - Veneto, Italy): LTM64/128 express, SD-128, Flexi. The majority of data were sampled with 512 Hz or 2048 Hz, but some patients had recordings with a sampling rate of 256 or 1024 Hz. Adtech electrodes (2008-mid 2019), PMT electrodes (since 2019) or Dixi electrodes (since mid 2018) were used.

### Preparatory steps to convert to BIDS

The BIDS specification defines a folder structure for storing different types of brain imaging and electrophysiology data and was recently extended to iEEG-BIDS^6^. The folder names convey information about the subject, session task and run and the user has to define this chain of *entities* (<key,value> pairs) to build the folder structure and name the files in an intuitive and BIDS-compatible manner.

In order to implement the iEEG-BIDS specification, different information needs to be extracted from the clinical source data. We identified 6 steps that were essential to organize clinical iEEG data in BIDS. These steps are: 1) assign a subject label, 2) define the session, task and run key-value pairs, 3) anonymize the data, 4) determine the resected brain area and label electrodes as resected, edge or non-resected, 5) annotate the binary files, and 6) convert to BIDS (see Figure 2 and 3).

**Figure 2.**
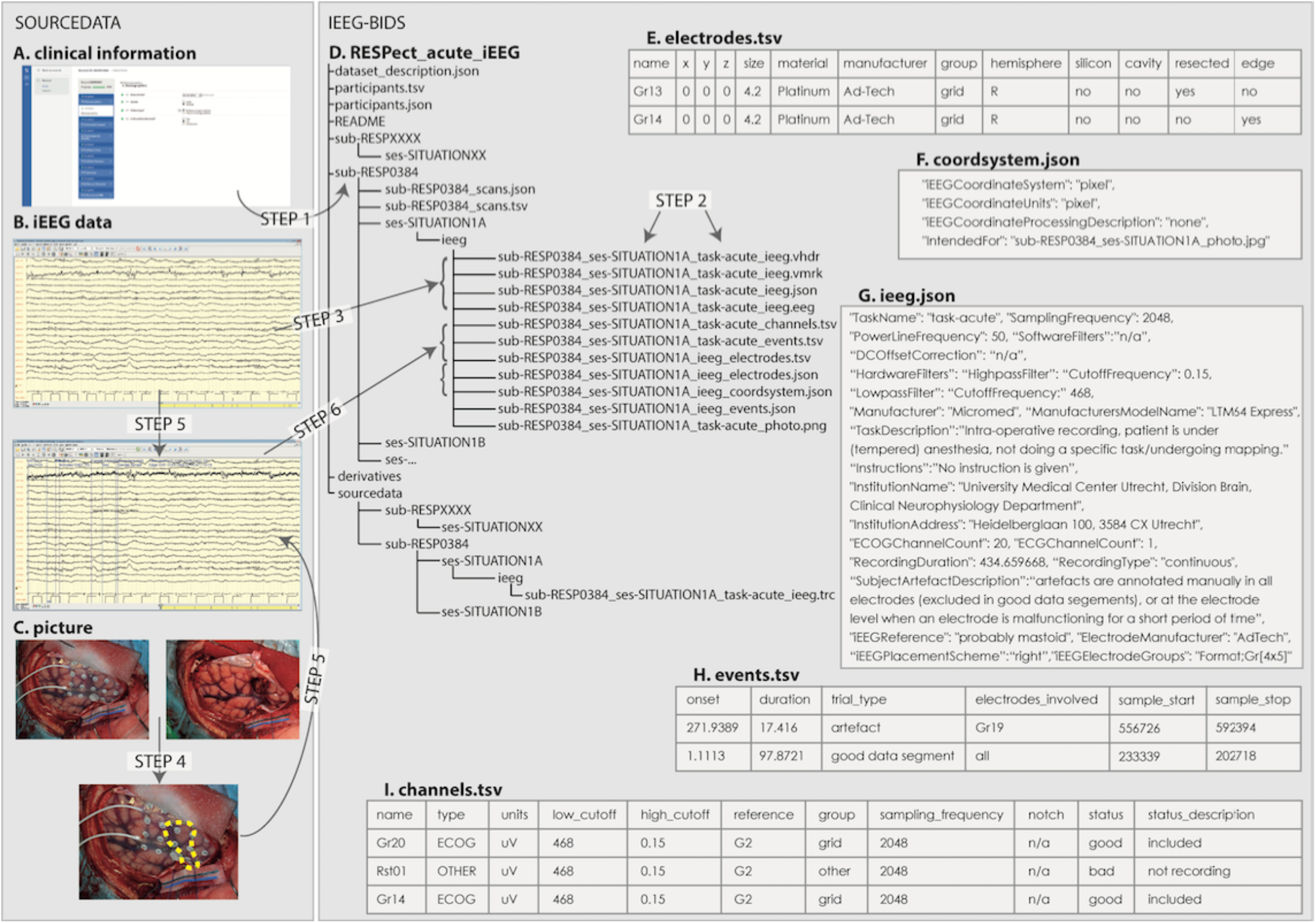
Overview of the steps required to convert the intraoperative ECoG recordings to iEEG-BIDS. In the left box, the sourcedata is displayed with A) the clinical information in an electronic data capture system, B) the raw (upper subplot) and annotated (lower subplot) acute ieeg recording in the clinical eeg system, C) the pictures showing the electrode positions: one pre-resection (left) and one post-resection (right), which are combined in a figure (below) with the resection indicated on top of the electrode grid with a dotted green line. In the right box, in D) the iEEG-BIDS data structure is displayed and in E-I) examples of BIDS specific files that should be present inside each sub-folder. The specific steps in this figure are explained in the text. All subplot results from subject RESP0384.

**Figure 3.**
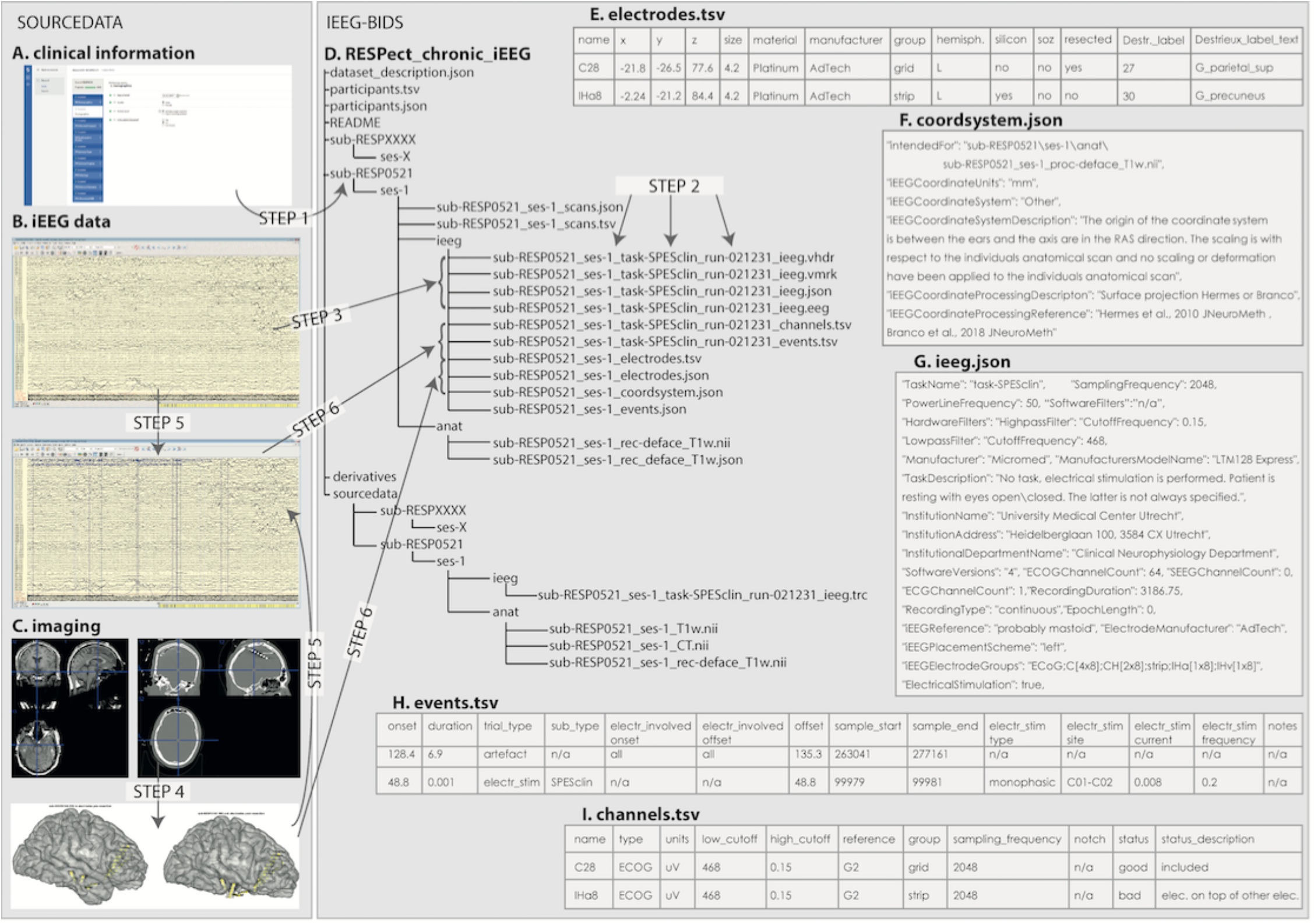
Overview of the steps and sourcedata required to convert the long-term iEEG recordings to iEEG-BIDS. In the left box, the sourcedata is displayed with A) the clinical information in an electronic data capture system, B) the raw (upper subplot) and annotated (lower subplot) long-term ieeg recording in the clinical eeg system, C) the defaced MRI (left) and coregistered CT (right), resulting in two patient specific brain renderings with the electrodes in yellow: one pre-resection and one post-resection. In the right box, in D) the iEEG-BIDS data structure is displayed and in E-I) examples of BIDS specific files that should be present inside each sub-folder. The specific steps in this figure are explained in the text. All subplots result from subject RESP0521, except subplot C which illustrates the imaging processes in SEEG subject RESP0749.

### Data Records

We constructed two separate RESPect iEEG-BIDS databases, one for intraoperative (see Figure 2) and one for long-term (see Figure 3) iEEG recordings. We here describe the steps performed to organize the clinical iEEG data in BIDS in detail.

### STEP 1: ASSIGN A SUBJECT LABEL

Parallel to the conversion of the iEEG data to iEEG-BIDS, we are putting clinical information like patient characteristics, epilepsy type, pathology and outcome after surgery of all patients included in RESPect in Castor, an electronic data capture system^13^. We use the same convention for subject labelling in the clinical and data part: the name should start with the RESP prefix and should be followed by a 4 digits number representing the code for a patient (e.g. RESPXXXX, where XXXX is a 4 digits number).

An overview of patients included in the RESPECT_acute_iEEG-BIDS and RESPECT_longterm_iEEG-BIDS is given in participants.tsv. This file contains the RESP-number, the number of sessions, the gender and the age of the patient when the data was recorded.

### STEP 2: DEFINE THE SESSION, TASK AND RUN KEY-VALUE PAIRS

The definition of the session, task and run differs between the two types of iEEG recordings and will be explained in the following subsections.

#### Intraoperative ECoG

We decided that each recording situation as explained in the Methods section, represents a *session* of the BIDS specification. We assigned the situation name to the key-value pair related to the *session* (e.g. ses-SITUATION1A). We did this, because the location of the electrodes changes with each recording situation, and are assigned at the session-level.

The intraoperative recordings we are currently converting to the BIDS format, are ongoing recordings during anesthesia without any stimulus (i.e. “resting state”). We decided to assign the label “acute” to the key-value pair related to the *task* (e.g. task-acute). Recordings where intraoperative somatosensory evoked potential (SSEP) is performed, or recordings where the patient is woken up to perform language or motor testing are defined as *task*-SSEP and *task*-stimulation.

We did not define the optional *run* key-value pair for intraoperative recordings, since only one run was recorded of each task. Once the session and task have been defined, it is possible to create the folder structure and name the files (Figure 2D).

#### Long-term iEEG

iEEG-files that were recorded within one monitoring period were categorized in the same *session*. When extra electrodes were added/removed during this period, the *session* was divided into ses-1a and ses-1b. Some patients had two long-term iEEG periods with, for example, first ECoG and second SEEG electrodes. These patients have a ses-1 and a ses-2. We use the optional *run* key-value pair to specify the day and the start time of the recording (e.g. run-021315, which means day 2 after implantation (day 1 of the monitoring period), at 13:15).

The *task* key-value pair in long-term iEEG recordings describes the patient’s state during the recording of this file. Different tasks have been defined, such as “rest” when a patient is awake but not doing a specific task, “sleep” when a patient is sleeping during the majority of the file, or “SPESclin” when the clinical SPES protocol was performed in this file. Other task definitions can be found in the annotation syntax (see step 5).

Once the session, run and task have been defined, it is possible to create the folder structure and name the files (Figure 3D).

### STEP 3: ANONYMIZE DATA

Intracranial EEG data are collected in (proprietary) binary formats that may include protected subject information. The binary format used in our center is by Micromed (TRC-file). We anonymized the TRC-files, because a BIDS data viewer is still missing and we wanted to allow our clinicians and researchers to visualize the anonymized and annotated data easily. We manually changed the patient names to RESPect identification number, the date of birth to 1-1-year in the Micromed patient identifier, and removed the patient names from recording montages. We subsequently ran Matlab code to further anonymize all fields in the rest of the TRC-file (see https://github.com/UMCU-EpiLAB/umcuEpi_acute_ieeg_respect_bids/anonymization and https://github.com/UMCU-EpiLAB/umcuEpi_longterm_ieeg_respect_bids/anonymization for the code implementation).

### STEP 4: DETERMINE THE RESECTED BRAIN AREA AND LABEL ELECTRODES AS RESECTED, EDGE OR NON-RESECTED

In both intraoperative and long-term iEEG recordings we added “resected”, “edge” and “cavity” labels to our <u>electrodes.tsv</u>, but the method used to do so differs (see description below).

#### Intraoperative ECoG

In intraoperative ECoG, we defined the “resected”, “edge” and “cavity” electrodes using the pictures taken in the operating room. We used a custom made-software^14^ to align the pre-resection and intermediate session pictures with the picture representing the end of the surgery. Then we drew the resection area on the post-resection picture and this was automatically projected on the pre-resection and intermediate session pictures (see Figure 1B, 2C; green/yellow dashed line). Electrodes that were completely or partly (so exactly on the edge) on top of the resected area were defined as resected. Electrodes that were partly on top of the resected area (so exactly on the edge) or within 0.5 cm of the edge of the resection were defined as edge. Electrodes that were above a resection cavity from an earlier surgery or a previous situation in the current surgery (so not recording brain signals) were defined as cavity.

#### Long-term iEEG

In long-term iEEG, we co-registered the pre-operative MRI to the CT with electrodes, and the post-operative MRI to the co-registered pre-operative MRI. We subsequently superimposed the CT with electrodes onto the co-registered post-operative MRI (see <u>ripts/elecPos03_process_postOperativeMRI.m</u>) and defined electrodes as “resected”, “edge” and “cavity” using the same definitions as above.

### STEP 5: ANNOTATE THE BINARY FILES WITH CUSTOM SYNTAX

In order to implement the BIDS specification, different metadata information is necessary (i.e. artefacts, good segment of the data, period of sleep etc.). We therefore decided to annotate our TRC-files with a custom syntax, using the proprietary Micromed visualization software (System Plus v. 1.04.0197) to include the metadata (Figure 2B and 3B). The syntax and scripts used to enrich the original trc-files and automatically create the BIDS files are available at https://github.com/UMCU-EpiLAB/umcuEpi_acute_ieeg_respect_bids/ for intraoperative and at https://github.com/UMCU-EpiLAB/umcuEpi_longterm_ieeg_respect_bids/ for long-term ECoG and SEEG recordings.

### STEP 6A: CONVERT TO BIDS - ELECTRODES AND COORDINATES

In the <u>electrodes.tsv</u>, the position, size and other properties of the iEEG contacts are stored (Figure 2E and 3E). The <u>coordsystem.json</u> file was intended for specification of the method and reference system used to determine the electrode positions (Figure 2F and 3F). The definition of the electrode positions differs between the intraoperative and long-term iEEG recordings and will be explained in the following subsections.

#### Intraoperative ECoG

Electrode coordinates during intra-operative recordings can be localized on 2D pictures taken during surgery. However, electrodes recording from the same brain tissue (i.e. overlapping parts from Situation 1A and 1B) would have different x,y coordinates based on different pictures taken during Situation 1A and 1B. The goal of these recordings is to identify epileptic versus normal tissue, and relate that to outcome. Therefore, we set the x, y, z coordinates in the electrodes.tsv file of the intraoperative iEEG data to zero, even though the iEEG-BIDS specification allows them to be given in 2D space from operative photos.

In the <u>coordsystem.json</u> file, we included the name to the picture taken before starting the recording (Figure 2C).

#### Long-term iEEG

The <u>electrodes.tsv</u> of long-term iEEG recordings contains the x, y, z coordinates, size and other properties of the electrodes. The CT was co-registered with the defaced T1 weighted pre-operative MRI. The MRI was segmented using Freesurfer software. The electrodes were localized on the CT-scan, corrected for brain shift and placed on the cortical surface (Figure 3C). The code (https://github.com/UMCU-EpiLAB/umcuEpi_longterm_ieeg_respect_bids/electrode_positions) to do this was adapted from Hermes et al.^15^ and Branco et al.^16^. In the <u>coordsystem.json</u>, the method and reference system used to determine the electrode positions is described. We additionally assigned electrodes to regions of the Destrieux^17^ and DKT atlases^18^ extracted using Freesurfer^19^.

### STEP 6B: CONVERT TO BIDS - INFORMATION ABOUT THE RECORDING AND CHANNELS USED

The <u>_ieeg.json</u> file contains metadata about the recordings (Figure 2G and 3G). In the field *iEEGElectrodeGroups*, we defined a way to express the used recording scheme. Specifically, we extracted the annotation we made in the TRC-file using the following syntax (see step 5): *Format;* followed by the electrode name and dimensions of the grid and/or strip/depth electrodes used.

In the example in Figure 2G, “Format;Gr[4×5]” implies that a rectangular grid with 20 electrodes was used for the intraoperative ECoG recording. In the example in Figure 3G, “ECoG;C[4×8];CH[2×8];strip;IHa[1×8];IHv[1×8]” implies that two grids and two strips were used for the long-term ECoG recording.

The <u>channels.tsv</u> file is intended for storing information related to the channels in a recording, such as the recording montage, sample frequency, units etc. (Figure 2I and 3I). The variables *status* and *status description* specify if the channels are available for usage and give a reason if a channel is not available. We used the annotations made in the TRC-file in step 5 to extract which channels contain good or bad signal, and defined the different reasons for bad signal in status description, for example:

1. Noisy - after visual inspection, a reviewer declared the channel as bad because the signal is noisy. These channels are annotated as “Bad;…” in step 5 (Figure 2B and 3B). The BIDS conversion will put their BIDS *status* to ‘bad’, with ‘noisy after visual inspection’ as BIDS *status description*.
2. Silicon - the electrode was placed on top of the silicon of another grid or strip, few brain signal is recorded. These channels are annotated as “Silicon;…”. The BIDS conversion will put their BIDS *status* to ‘bad’, with ‘electrode on top of other electrode’ as BIDS *status description*.
3. Screw - this annotation was only present in SEEG recordings. It defines an electrode that was not recording cortical signals, but located in the screw outside the brain. This was determined from the electrodes extracted from the CT and co-registered on the pre-operative MRI. These channels were annotated as “Screw;…”. The BIDS conversion will put their BIDS *status* to ‘bad’, with ‘located in screw’ as BIDS *status description*.

### STEP 6C: CONVERT TO BIDS - EVENTS IN THE RECORDING

The <u>events.tsv</u> file contains a table with the onset, duration, and channels involved in events present in a recording. We annotated the onset and offset of events in the TRC-files with a specific syntax in step 5. These annotations were converted to onset and duration in the events.tsv files. The events differed between intraoperative and long-term iEEG recordings and were explained in more detail in the <u>sub-RESPXXXX_ses-X_events.json</u> file in the subject’s directory of the respective iEEG-BIDS database.

#### Intraoperative ECoG

In intraoperative ECoG, we used *event* definitions to mark good, clean data segments without artefacts due to equipment in the operating room or due to surgical manipulation, and without burst-suppression as a result of remnants of propofol anesthesia. If intra-operative SSEP was performed or if the patient was woken up to perform language or motor testing, additional *event* annotations were added in a column defined in an accompanying _events.json.

#### Long-term iEEG

In long-term iEEG, *task* definitions and *event* annotations are often coupled: if a task (for example sleep) was defined and annotated at the beginning of the file in step 5, a period of sleep was annotated in the file with Sl_on and Sl_off. This period of sleep was stored in the <u>events.tsv</u> as an event with onset (e.g. time corresponding to SI_on marker) and duration (time between SI_on and SI_off markers). Artefacts, seizures, stimulation, motor tasks, etc. are also annotated and added in the <u>events.tsv</u>.

The optional <u>scans.tsv</u> file contains an overview of all the files present in a session of a patient, and the type of tasks and events present in these files. This is useful to decide which recordings can be used to answer a specific research question.

### STEP 6D: CONVERT TO BIDS - TRC TO SUPPORTED FILE FORMAT

TRC-files are not part of the set of supported binary file formats of the BIDS specification. We therefore converted our iEEG data to BrainVision Core Data Format (.vhdr, .eeg, .vmrk).

### STEP 6E: CONVERT TO BIDS - STRUCTURE SOURCE DATA

The anonymized and annotated TRC-files of each patient were stored in their subfolder in the sourcedata folder. For intraoperative iEEG, this folder also contains the original pictures of the electrode positions taken in the operating room (before aligning them with the post-resection image and drawing the resection cavity). For long-term iEEG, this folder also contains CT scans with electrode positions and raw T1 weighted MRI scans. The defaced MRI is located in the anat-subfolder in each specific patient folder. The derivatives-folder contains a freesurfer folder with each subject’s MRI scans processed with freesurfer^19^.

The data of six intraoperative, and a sleep recording, a recording containing a seizure and (if available) a recording containing a stimulation session of three long-term ECoG and three long-term SEEG patients was converted to the iEEG-BIDS format as described above and are stored in openneuro.org with the following doi: 10.18112/openneuro.ds003400.v1.0.0, 10.18112/openneuro.ds003399.v1.0.0.

#### Technical Validation

We ran our example patients through the BIDS Validator App^20^. The long-term ECoG and SEEG examples passed the validation procedure. The intraoperative ECoG examples passed the validation procedure with zero’s as electrode coordinates in the electrodes.tsv (details step 6a).

## Data Availability

data availability at openneuro.org with the following doi: 10.18112/openneuro.ds003400.v1.0.0, 10.18112/openneuro.ds003399.v1.0.0.

## Code Availability

Intraoperative ECoG: https://github.com/UMCU-EpiLAB/umcuEpi_acute_ieeg_respect_bidsfor intraoperative ECoG and https://github.com/UMCU-EpiLAB/umcuEpi_longterm_ieeg_respect_bids/for long-term iEEG.

## Acknowledgements

We would like to thank the members of the RESPect group for gathering the data in a complete and consistent matter. A special thanks to Janine Ophorst and Anouk Velders for their help with respect to gathering informed consent from the prospective patients. We thank the students and research assistants who annotated part of the data: Jaap van der Aar, Giulio Castegnaro, Daniel Groothuysen, Emile d’Angremont, Merel Wassenaar, Paul Smits, Bram Knipscheer, Sifra Blok.

WZ was supported by the Alexandre Suerman Stipendium 2015.

MD was supported by the grant LSHM16054-SGF.

MZ was supported by the ERC starting grant 803880

DvB was supported by NEF 17-07.

DH was supported by NIH grant R01 MH122258-01.

## Author contributions

MD, DvB and WZ contributed equally in building the pipeline, preparing the data and writing the protocol, scripts and paper. DH critically reviewed our pipeline, shared her code for determining the electrode position of the long-term ECoG and SEEG electrodes, and revised the paper. FL and MZ supervised the process and revised the paper. The members of the RESPect group all contributed in a way to gather or process the data.

